# Serial neoGFAP^™^ outperforms total GFAP for monitoring and 6-month outcome discrimination after moderate to severe traumatic brain injury: an exploratory single-site cohort study

**DOI:** 10.64898/2026.09.01.26361862

**Authors:** Kevin K. Wang, Guangzheng Cai, Khadija Boukholda, Firas Kobeissy, Eman Elbayoumi, Devin Jackson, Katie Tehas, Kristy Radeker, Anthony DeLizza, Caroline Popper, Spyridoula Tsetsou, Claudia Robertson, William E. Haskins

## Abstract

**Background:** Serial glial fibrillary acidic protein (GFAP) trajectories have become an important framework for contextualizing evolving secondary-injury pathophysiology after moderate-to-severe traumatic brain injury (msTBI). However, total GFAP pools release and clearance signals that may be less useful for longitudinal bedside decisions than a proteoform-resolved assay. We compared total GFAP with neoGFAP™, defined here as calpain-generated GFAP proteoforms intended to index active astroglial proteolysis during the subacute phase.

**Methods:** We analyzed 651 serial serum samples from 95 msTBI patients from a previously described single-site cohort. Total GFAP and neoGFAP™ were measured on the same MSD platform from 6 to 240 hours after injury. Early (6–72 h) and late (96–240 h) windows, data-derived tertiles, and serial trajectory summaries were calculated directly from serial samples. Models were benchmarked against age plus admission post-resuscitation Glasgow Coma Scale (GCS) and the admission IMPACT extended risk score using five-fold stratified cross-validation. Outcomes were unfavorable outcome (GOSE 1–4), less-than-good recovery (GOSE 1–6), Disability Rating Scale (DRS) ≥15, mortality, and neuroimaging worsening at 6 months.

**Results:** The cohort contributed 95 serial biomarker profiles, with 90 participants evaluable for 6-month GOSE and 89 for DRS. Unfavorable outcome occurred in 57/90 (63.3%), and less-than-good recovery in 79/90 (87.8%). For unfavorable outcome, IMPACT plus early neoGFAP™ reached AUROC 0.85 versus 0.84 for IMPACT plus early total GFAP and 0.81 for IMPACT alone. For less-than-good recovery, IMPACT plus late neoGFAP™ achieved AUROC 0.90 versus 0.84 for late total GFAP and 0.82 for IMPACT alone. Secondary analyses for DRS, mortality, and neuroimaging worsening showed smaller differences.

**Conclusions:** In this retrospective analysis, neoGFAP™ provided clearer incremental value than total GFAP for recovery-oriented monitoring, especially when late-window reassessment of patients who remained at risk for less-than-good recovery was required. Results support prospective testing of neoGFAP™ as a pathophysiology-informed adjunct to serial bedside decision making, repeat-assessment thresholds, and recovery stratification.

## Introduction

Adult moderate-to-severe traumatic brain injury (msTBI) evolves over days rather than minutes. Physicians frequently rely on repeated neurological examinations and, in higher-risk scenarios, repeat computed tomography (CT) imaging to monitor progression or resolution of intracranial pathology. Yet serial examinations are often confounded by intubation, sedation, decompressive surgery, or hemodynamic instability, and repeat imaging adds transport, workflow, and resource burdens. A useful blood biomarker in this setting should help distinguish between a patient whose astroglial injury burden is biologically settling from one whose risk remains persistently high. Total GFAP has become clinically important because it rises early after injury and tracks astroglial damage, but it remains an aggregate signal that can reflect structural release, redistribution, proteolysis, and clearance rather than a single biological process.

msTBI is commonly operationalized as TBI with profound early neurological dysfunction, often corresponding to a post-resuscitation Glasgow Coma Scale (GCS) score of 3–8, and is better understood as a prolonged pathophysiologic syndrome than as a single mechanical event. In the United States, the Centers for Disease Control and Prevention (CDC) reports more than 69,000 TBI-related deaths in 2021 [1], while globally the World Health Organization (WHO) attributes roughly 10% of years lived with disability to injuries [2,3]. Long-term disability is common: 57% of msTBI survivors alive at 5 years are moderately or severely disabled and 55% are unemployed [1,4]. This underscores why monitoring tools that remain informative across ICU management, rehabilitation, and long-term recovery are clinically important.

This distinction matters for monitoring because the dominant clinical need is not merely to identify a statistically significant between-group difference; it is to follow evolving pathophysiology within an individual as injury progresses from acute neurocritical care toward rehabilitation and longer-term recovery. Total GFAP is informative for injury burden, yet its value for serial bedside decision making may be limited when clinicians want to know whether a patient is simply clearing an early injury signal or whether secondary-injury biology remains active. The broader GFAP proteoform literature also suggests that assay performance can vary because different platforms capture different mixtures of intact and fragmented proteoforms [5].

We define **neoGFAP™** as mechanistically anchored GFAP species of 38–40 kDa bearing a defined calpain-cleaved neo-N terminus and heterogeneous C-termini, released from injured astrocytes into the extracellular fluid. Following TBI, excitotoxic and calcium-dependent cascades activate calpains, which cleave astrocytic GFAP and generate proteoforms with cleavage-dependent neo-epitopes that are not present on intact GFAP [6]. Because neoGFAP™ is generated by injury-associated proteolysis rather than by bulk release alone, it may more directly index astrocyte structural injury and debris processing than total GFAP. In a monitoring setting, this raises a clinically relevant possibility: serial neoGFAP™ could be more helpful to physicians than total GFAP when used to follow within-patient trajectory, support recovery stratification, and inform the threshold for continued close observation or reassessment.

Prior work provides trajectory context for interpreting these data. Robertson et al. showed that serial serum GFAP trajectories separated patients into low, middle, and high groups associated with greater intracranial pressure (ICP) burden, lower cerebral perfusion pressure (CPP) and brain tissue oxygenation (PbtO_2_), higher mortality, and worse 6-month outcome [7]. Wang et al. showed that axonal markers neurofilament light chain (NfL) and phosphorylated neurofilament heavy chain (pNfH) rise more slowly and peak later after injury [8]. Together, these studies show that biomarker time courses can add clinically meaningful information beyond a single admission value. The present study asks whether a proteoform-resolved astroglial marker can strengthen this serial-monitoring paradigm by outperforming total GFAP in the outcomes most relevant to longitudinal clinical management.

**Schematic 1.**
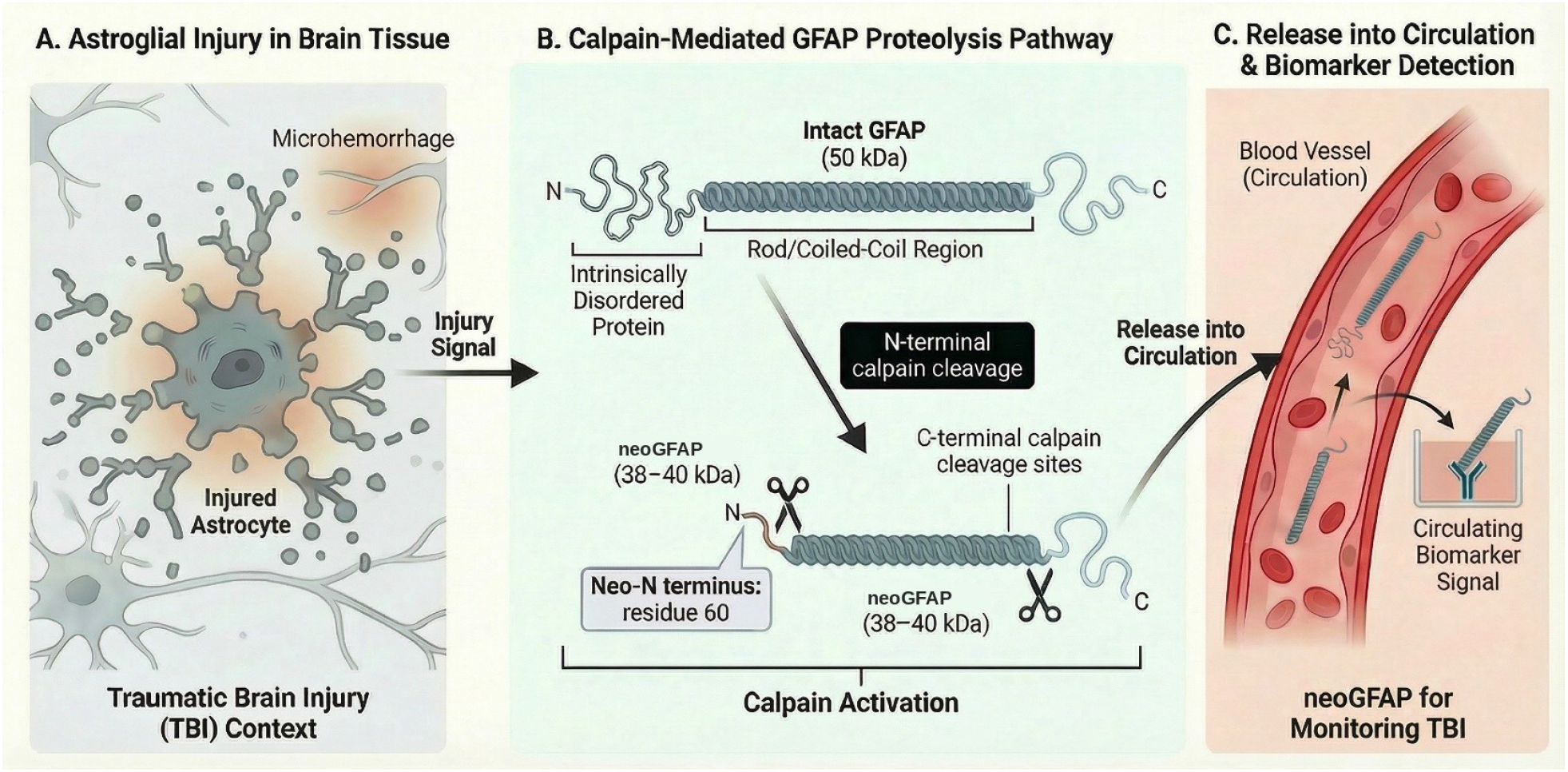
Mechanistic basis of neoGFAP™ generation and release into circulation. (A) Astroglial injury within the TBI context, showing an injured astrocyte in association with tissue damage features such as microhemorrhage. (B) Calpain-mediated GFAP proteolysis: intact GFAP contains an N-terminal intrinsically disordered protein (IDP) region, the rod/coiled-coil region, and a C-terminal IDP region; N-terminal calpain cleavage exposes a neo-N terminus at residue 60, and additional C-terminal cleavage generates predominantly 38–40 kDa neoGFAP™ fragments. (C) Release of neoGFAP™ proteolytic fragments into the bloodstream for biomarker detection.

## Methods

### Study cohort and data sources

This was a retrospective analysis of archived serum samples from patients with msTBI. Serial biomarker data comprised 651 draw-level serum measurements from 95 participants across the first 10 post-injury days. De-identified demographics, admission injury characteristics, IMPACT extended-model risk scores (age, motor GCS category, pupillary reactivity, hypoxia, hypotension, Marshall CT classification, traumatic subarachnoid hemorrhage, epidural hematoma, glucose, and hemoglobin), Marshall CT classifications, delayed-hematoma variables, adverse events, and 6-month GOSE and DRS outcomes were provided in the parent clinical dataset.

### Biomarker processing

Serum total GFAP and neoGFAP™ were measured on the same Meso Scale Discovery (MSD) platform using in-house proprietary capture/detection monoclonal antibody pairs designed to distinguish total GFAP from a cleavage-defined neo-N-terminal epitope. NfL was not measured in this cohort due to sample volume limitations. Early and late windows were defined a priori as 6–72 h and 96–240 h after injury. For each analyte we calculated window means, medians, sample counts, and whole-window slopes from the raw serial data. Trajectory visualization and tertile grouping were computed de novo from the raw data rather than imported from earlier analyses. Total GFAP and neoGFAP™ were measured on the same MSD platform, and all AUROC comparisons in this manuscript are within-platform.

### Outcome definitions

Unfavorable outcome was defined as 6-month GOSE 1–4 versus 5–8. Less-than-good recovery was defined as GOSE 1–6 versus 7–8. DRS ≥15 was used to capture severe disability or death. Mortality was defined as 6-month GOSE of 1. CT worsening was defined from the difference between initial and worst Marshall CT category. The hemorrhagic neuroworsening composite combined CT worsening, delayed hematoma, and hemorrhagic CNS adverse events. Delayed hematoma type and start day were additionally summarized descriptively. Hourly ICU GCS was not used as a primary neuroworsening definition because retrospective interpretation is heavily confounded by clinical factors such as intubation and sedation.

### Statistical analysis

Biomarker distributions were compared non-parametrically. Prediction was evaluated using five-fold stratified cross-validation with pooled out-of-fold predictions. Benchmarks included age plus admission GCS and admission IMPACT extended risk scores. We report AUROC (2 decimals), area under the precision-recall curve (AUPRC), and Brier score. Simple bedside summaries included early and late window means plus early tertile and quartile groupings; richer trajectory summaries were recalculated directly from the raw serial data and are presented in the Supplementary Appendix. Analyses were performed in Python 3 (scikit-learn ≥1.3).

## Results

### Cohort profile

The serial biomarker cohort included 95 participants, of whom 90 had 6-month GOSE and 89 had 6- month DRS. The cohort was young and severely injured, with mean age 33.2 years, 81.1% male sex, median admission GCS 7, and median IMPACT risk of unfavorable outcome 0.43. This clinical-severity backdrop is important because it frames the added question: whether serial neoGFAP™ improves both total GFAP and established clinical prognostication when physicians are reassessing patients over time. Cohort characteristics and endpoint frequencies are summarized in Table 1.

**Table 1.** Cohort characteristics and 6-month endpoint frequencies.

| Characteristic | Value |
| --- | --- |
| Participants with serial biomarker data | 95 |
| Participants with 6-month GOSE | 90 |
| Participants with 6-month DRS | 89 |
| Age, years (mean ± SD) | 33.2 ± 13.9 |
| Male sex | 77/95 (81.1%) |
| Admission GCS sum, median [IQR] | 7 [4, 8] |
| IMPACT risk of unfavorable outcome, median [IQR] | 0.43 [0.20, 0.64] |
| Unfavorable outcome (GOSE 1–4) at 6 months | 57/90 (63.3%) |
| Less-than-good recovery (GOSE 1–6) at 6 months | 79/90 (87.8%) |
| DRS $\geq 15$ at 6 months | 25/89 (28.1%) |
| 6-month mortality | 14/90 (15.6%) |
| CT worsening | 15/95 (15.8%) |
| Delayed hematoma | 30/95 (31.6%) |
| Hemorrhagic neuroworsening composite | 37/95 (38.9%) |

### Serial trajectories and data-derived risk grouping

Both analytes generally declined over the first 10 post-injury days as expected, but serial separation remained visible when participants were grouped by analyte-specific early tertiles (Figure 1). That pattern is clinically important because it mirrors the central insight of the previous GFAP trajectory paper [7] while showing that the neoGFAP™ gradient is sharper for recovery-oriented outcomes: participants in the highest early neoGFAP™ tertile remained biologically high-risk across the monitoring window despite time since injury.

**Figure 1.**
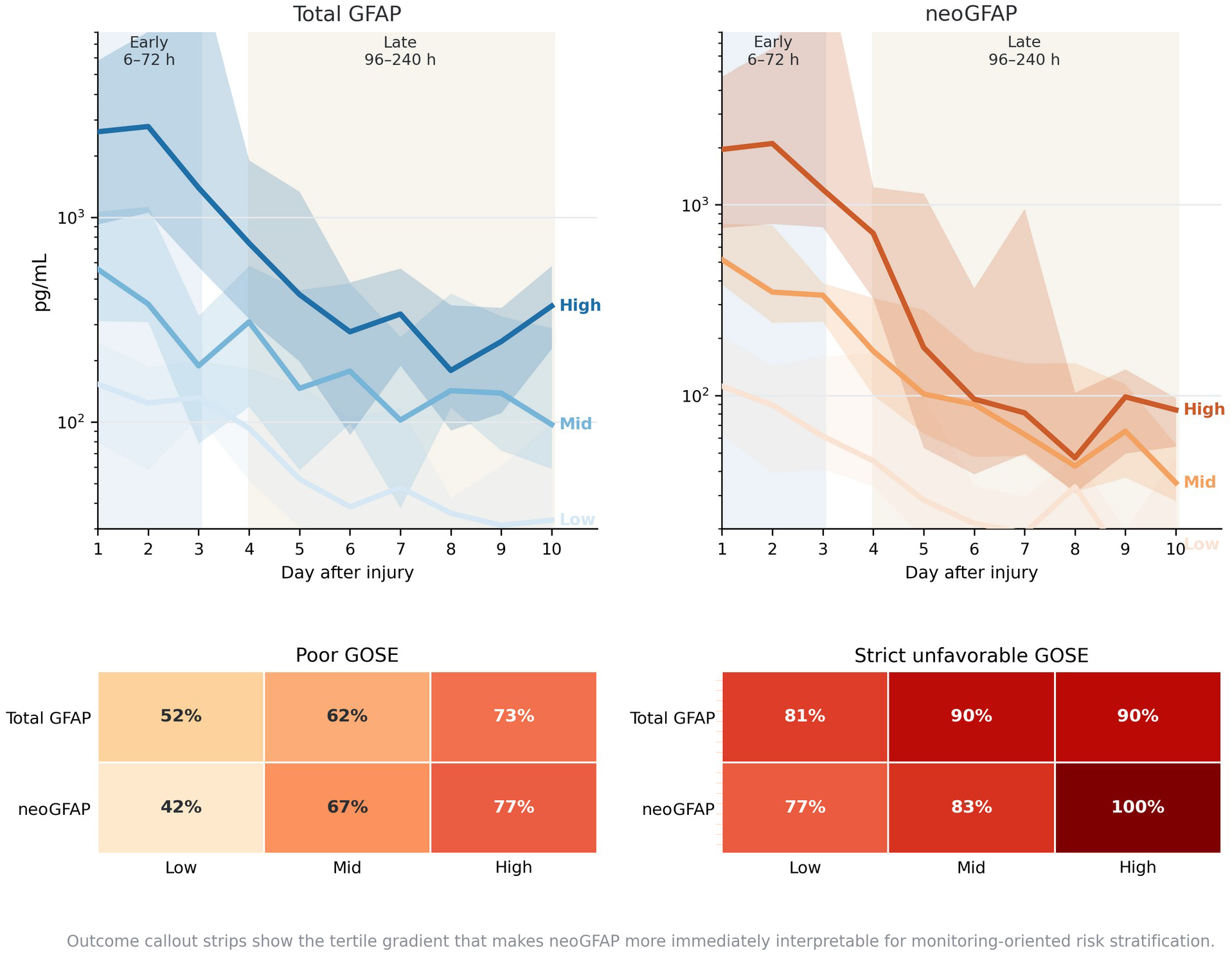
Serial total GFAP (left) and neoGFAP™ (right) trajectories stratified by analyte-specific early tertiles. Shaded vertical bands mark the early (6–72 h) and late (96–240 h) monitoring windows. Thick lines show tertile medians and translucent ribbons show the interquartile range. Outcome-callout strips beneath the trajectories summarize unfavorable outcome and less-than-good recovery rates across early tertiles.

The highest early neoGFAP™ tertile showed the clearest downstream gradient: 76.7% unfavorable outcome, 100.0% less-than-good recovery, 40.0% DRS ≥15, and 26.7% mortality (Table 2). Figure 1 also shows the corresponding early total-GFAP gradient, which was present but less sharply separated for less-than-good recovery. This pattern is consistent with the argument that early neoGFAP™ may provide physicians with a simpler and more interpretable monitoring-oriented risk framework.

**Table 2.** Early neoGFAP™ tertiles and downstream 6-month outcomes.

| Early tertile | N | Unfavorable outcome | Less-than-good recovery | DRS ≥15 | Mortality |
| --- | --- | --- | --- | --- | --- |
| Low | 30 | 11/26 (42.3%) | 20/26 (76.9%) | 3/26 (11.5%) | 2/26 (7.7%) |
| Mid | 30 | 20/30 (66.7%) | 25/30 (83.3%) | 10/29 (34.5%) | 4/30 (13.3%) |
| High | 30 | 23/30 (76.7%) | 30/30 (100.0%) | 12/30 (40.0%) | 8/30 (26.7%) |

*Tertiles were derived directly from the 6–72 h neoGFAP™ window means. Denominators reflect outcome-evaluable subjects within each tertile.*

### Functional outcome discrimination

neoGFAP™ outperformed total GFAP most consistently for the outcomes most relevant to longitudinal clinical management. For unfavorable outcome, early neoGFAP™ was already stronger than early total GFAP at the distribution level, with higher early values in participants with poor outcome and a modest AUROC advantage once added to IMPACT. For less-than-good recovery, the late window provided the clearest separation, supporting the idea that proteolytic GFAP fragments may better capture persistent secondary-injury biology during the phase when clinicians are considering whether a patient is stabilizing, plateauing, or remaining biologically high-risk (Figure 2).

**Figure 2.**
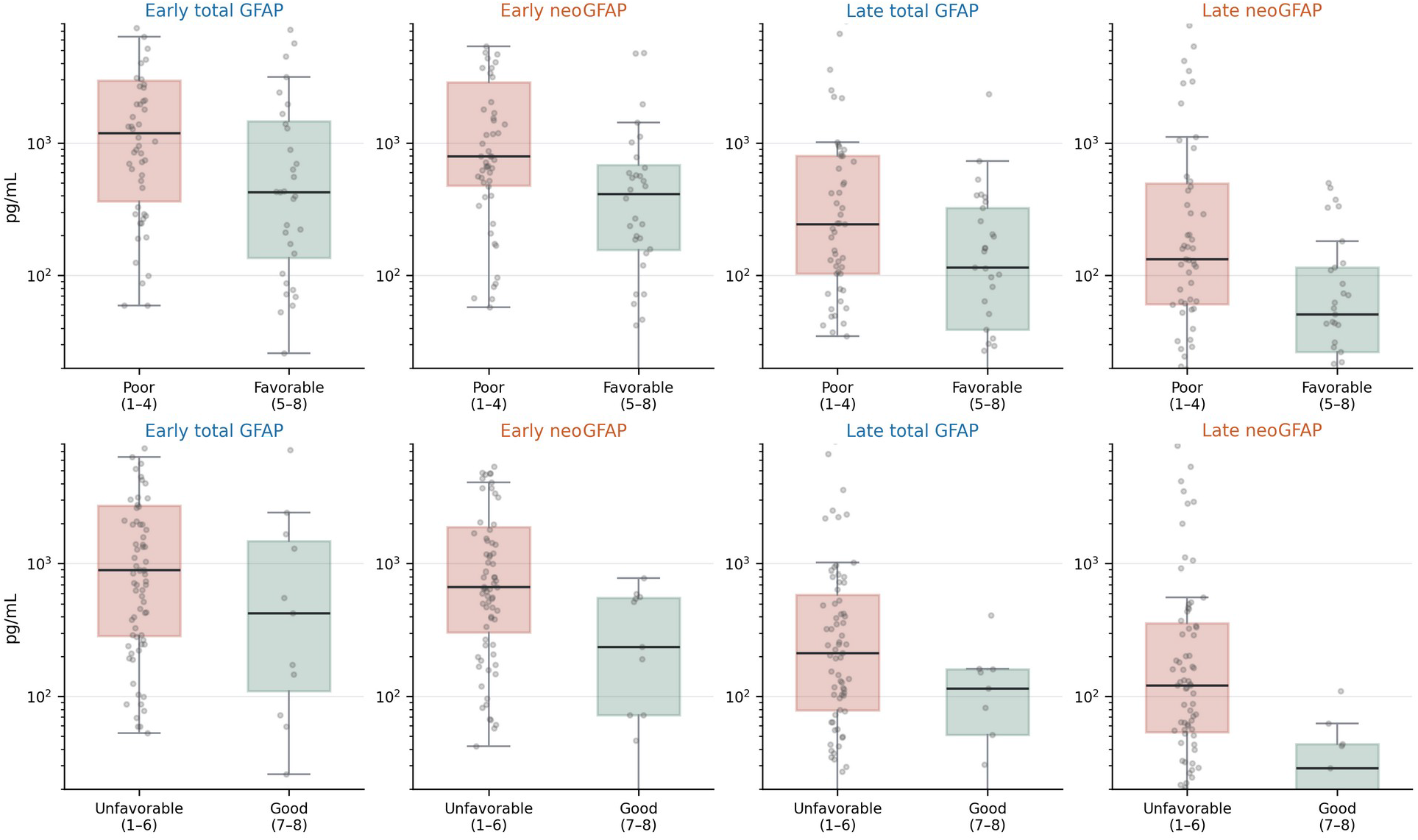
Early and late total GFAP and neoGFAP™ distributions across unfavorable versus favorable and less-than-good versus good 6-month GOSE. Top row: unfavorable outcome (GOSE 1–4) versus favorable (GOSE 5–8). Bottom row: less-than-good recovery (GOSE 1–6) versus good recovery (GOSE 7–8). Boxplots are overlaid on individual subject means and displayed on a logarithmic scale.

Cross-validated models reinforced this pattern. IMPACT plus early neoGFAP™ yielded AUROC 0.85 for unfavorable outcome, compared with 0.84 for IMPACT plus early total GFAP and 0.81 for IMPACT alone. For less-than-good recovery, IMPACT plus late neoGFAP™ reached AUROC 0.90, compared with 0.84 for late total GFAP and 0.82 for IMPACT alone. Because the central clinician-facing question was whether neoGFAP™ improved serial outcome monitoring beyond total GFAP, the manuscript emphasizes these head-to-head functional comparisons; the primary clinician-relevant model performance is summarized in Table 3, and fuller secondary endpoint grids are provided in the Supplementary Appendix.

**Table 3.**
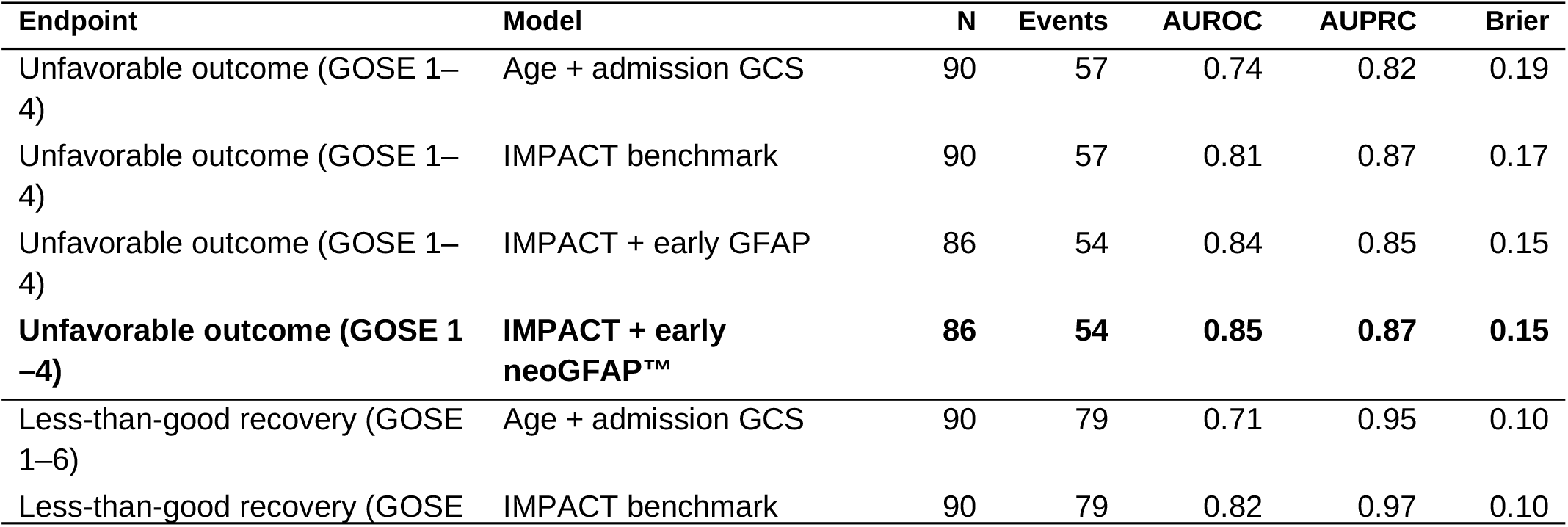
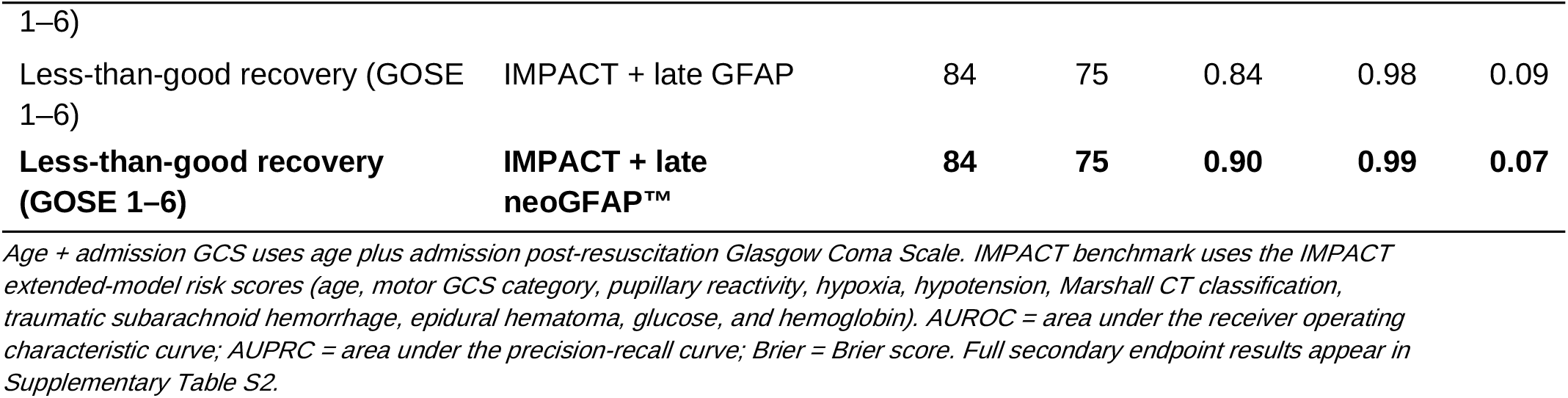
Cross-validated performance of primary clinician-relevant models for unfavorable outcome and less-than-good recovery at 6 months.

### Secondary outcomes

Secondary analyses are reported in the Supplementary Appendix. DRS and mortality showed smaller differences between neoGFAP™ and total GFAP than the GOSE analyses, and cross-validated discrimination for CT worsening, delayed hematoma, and the hemorrhagic neuroworsening composite remained weak (Supplementary Table S2). This contrasted with Robertson et al. [7], whose earlier total-GFAP trajectory paper emphasized strong descriptive links between high GFAP trajectories and hemorrhagic complications. In our analogous descriptive reanalysis using early neoGFAP™ tertiles, the high tertile still showed more severe Marshall worsening (≥+2 category: 5/30 [16.7%] vs 1/30 [3.3%] in the low tertile) and more delayed hematoma (12/30 [40.0%] vs 7/30 [23.3%]), but delayed hematoma start day was usually day 1 and the signal did not translate into robust predictive AUROCs (Supplementary Table S7; Supplementary Figure S3). An exploratory Robertson-style latent-class analogue reached a similar conclusion (Supplementary Table S8; Supplementary Figures S4–S5), with somewhat cleaner class separation for neoGFAP™ than for MSD total GFAP but no reproduction of Robertson’s strong mortality gradient. These patterns suggest that the clearest practical value of neoGFAP™ in this dataset lies in recovery-oriented monitoring rather than acting as a stand-alone surrogate for every downstream complication.

### Comparison with prior trajectory papers

Prior serial-biomarker studies in TBI differ in analyte, sampling horizon, and analytic framework, so the most useful comparison is conceptual rather than literal platform matching. Within that broader context, the present findings align partially, but not completely, with Robertson et al., who used Quanterix SIMOA total GFAP in a 10-day msTBI cohort and identified three trajectory classes; the high trajectory group was associated with 26.9% mortality, 88.5% unfavorable 6-month outcome, greater intracranial hypertension, lower CPP and PbtO_2_, and delayed hematoma in 69.2% of cases [7]. Wang et al. showed that NfL and pNfH rise more gradually and peak later, reaching maximal concentrations around 20–30 days [8]. Our primary MSD-based study differs in design: we recalculated early and late summaries plus data-derived tertiles directly from the raw serial data and tested cross-validated endpoint prediction rather than relying on latent class assignment. To bridge those frameworks, we ran an exploratory Robertson-style latent-class analogue on the daily-binned MSD data (Supplementary Table S8; Supplementary Figures S4–S5). Both analytes preferred four classes by BIC, but the three-class solutions retained high posterior probabilities and were used for apples-to-apples comparison with Robertson’s published three-class model. MSD total GFAP partially resembled the published SIMOA result for unfavorable outcome (high class 87.5%), whereas neoGFAP™ showed somewhat cleaner separation for unfavorable outcome and ICU-burden-type measures. Selected model comparisons are shown in Figure 3.

**Figure 3.**
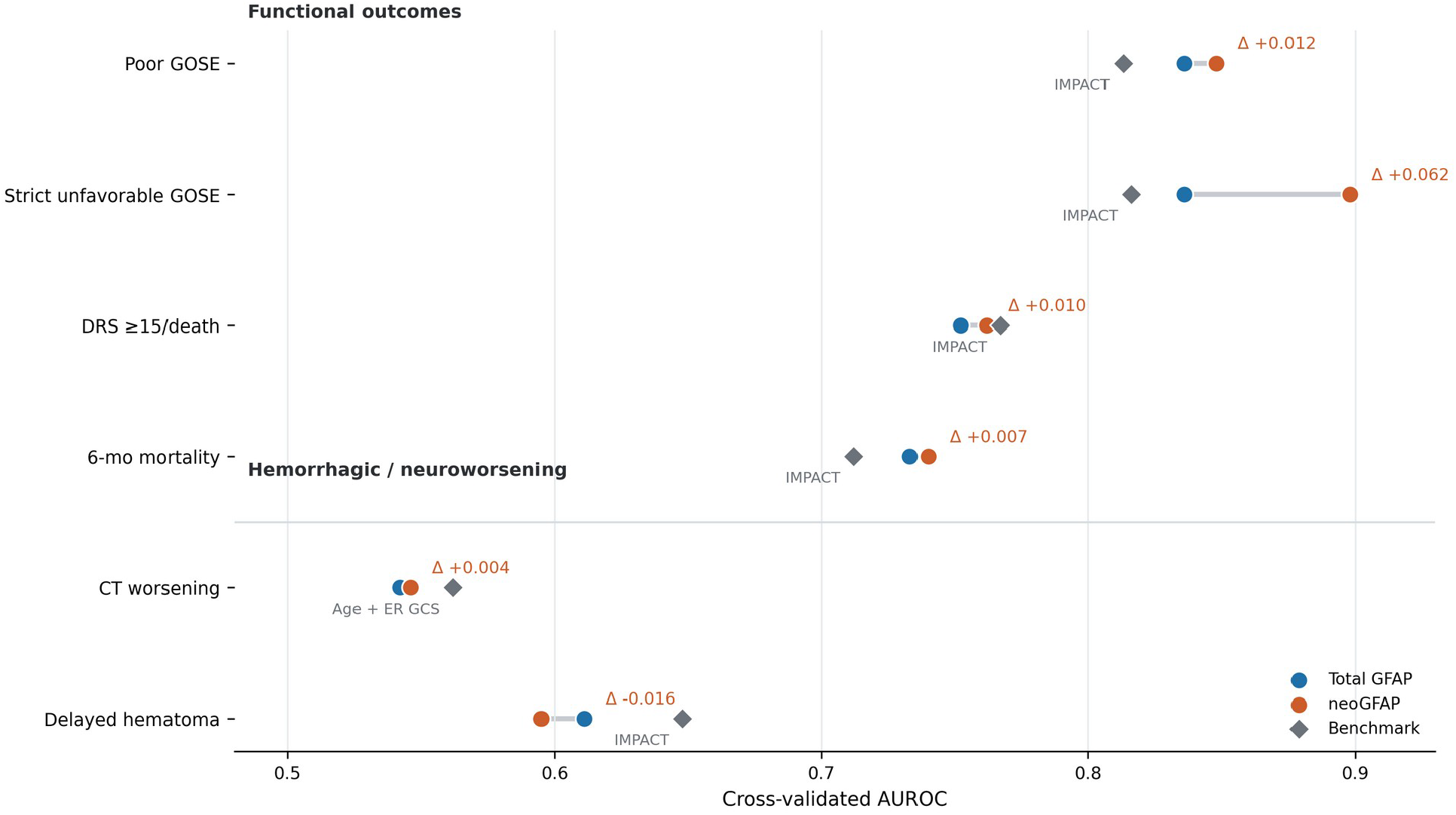
Benchmark-versus-biomarker dumbbell plot for selected functional and neuroworsening endpoints. Gray diamonds show the benchmark model (IMPACT or age + admission GCS), blue circles show matched total-GFAP models, and orange circles show matched neoGFAP™ models. Farther-right orange points for unfavorable outcome and less-than-good recovery highlight the clearest clinician-relevant advantage of neoGFAP™.

## Discussion

This retrospective analysis supports a focused clinical message: neoGFAP™ may be more helpful to physicians than total GFAP when the practical task is serial monitoring and recovery-oriented decision support after msTBI. The strongest signal did not come from increasingly complex modelling alone, but from a consistent head-to-head pattern in which early risk grouping and late neoGFAP™ burden outperformed matched total-GFAP summaries for functional outcomes, especially less-than-good recovery (GOSE 1–6) but also unfavorable outcome (GOSE 1–4).

That distinction matters clinically because total GFAP and neoGFAP™ are not answering exactly the same question. Total GFAP remains a useful astroglial injury marker, but neoGFAP™ is intended to capture linkage to injury-associated proteolysis. The main clinical need is not simply to demonstrate a between-group difference; it is to provide a biomarker that changes meaningfully within an individual as injury evolves. In this dataset, early risk grouping and late neoGFAP™ burden were more informative than matched total-GFAP summaries when physicians would be deciding how intensively to continue monitoring, whether a patient still warranted close observation, or whether recovery appeared biologically more plausible despite an examination that remained limited.

From a translational perspective, the promise is therefore a trajectory-aware interpretation rather than a single universal threshold. A monitoring biomarker is only as useful as its ability to support repeated interpretation over time. Figure 4 summarizes the clinician-facing logic suggested by these data: an early neoGFAP™ tertile establishes initial risk context, late-window persistence refines whether biology appears to be settling or remaining active, and serial change supports continued close observation, repeat assessment, and recovery stratification. If validated prospectively, serial neoGFAP™ could complement neurological examination and imaging by providing longitudinal context for continued ICU observation, repeat-imaging thresholds, step-down planning, and risk communication with families.

**Figure 4.**
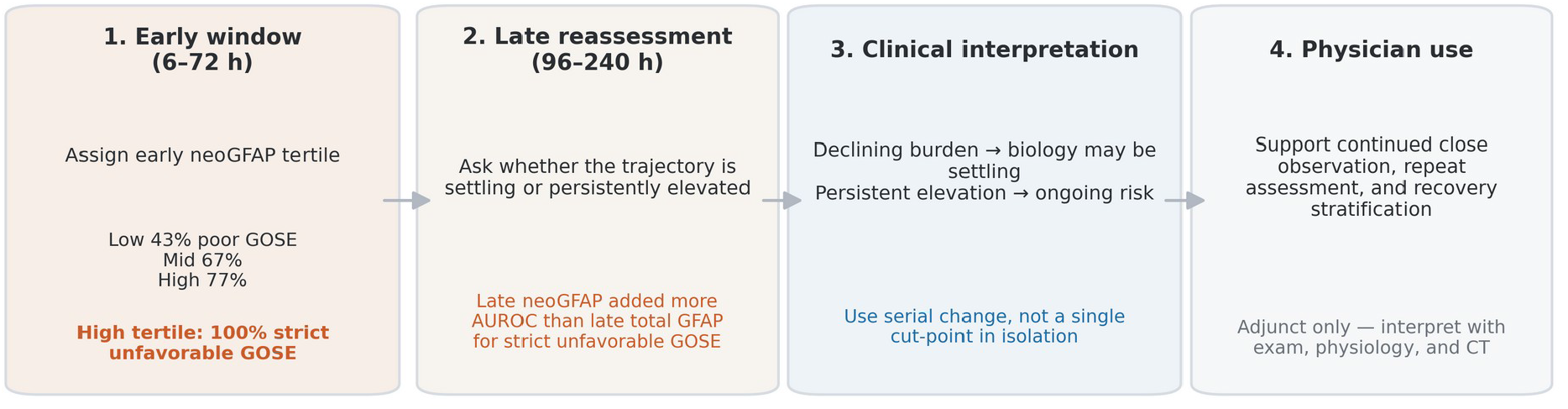
Clinician-facing monitoring framework suggested by the current data. An early neoGFAP™ tertile establishes initial risk context; late-window persistence versus decline refines interpretation; serial change is then used as an adjunct to support continued close observation, repeat assessment, and recovery stratification. Conceptual aid only, not a validated standalone decision rule.

To contextualize these AUROCs against the established prognostic benchmark for moderate-to-severe TBI, the IMPACT prognostic models derived by Steyerberg et al. reported cross-validated AUROC of ≈0.78 for both 6-month mortality and unfavorable outcome with the Core model (age + motor GCS + pupillary reactivity), rising to ≈0.80–0.83 for the Extended model that adds Marshall CT classification, traumatic subarachnoid hemorrhage, epidural hematoma, hypoxia, and hypotension, and to ≈0.83–0.87 for the Lab model that further adds admission glucose and hemoglobin [18]. Subsequent prospective independent validation reported AUROC 0.78 (mortality) and 0.76 (unfavorable) for Core, and 0.83 and 0.79 for Extended [19]. In our Baylor cohort the IMPACT-extended benchmark reached AUROC 0.81 for unfavorable outcome and 0.82 for less-than-good recovery — in the middle of the published Extended-model range — confirming that this cohort behaves like a typical Extended-benchmarked msTBI sample and provides a well-calibrated anchor for evaluating incremental biomarker value. Against that anchor, adding early neoGFAP™ to IMPACT raised AUROC to 0.85 for unfavorable outcome, and adding late neoGFAP™ raised AUROC to 0.90 for less-than-good recovery. These gains are notable because IMPACT-extended is already near the top of the published discrimination range for this endpoint, so the biomarker increment reflects genuinely orthogonal information rather than compensation for a weak clinical baseline. Because IMPACT is a static admission-time predictor while serial neoGFAP™ is inherently temporal, the two are complementary: IMPACT establishes initial risk context, and serial neoGFAP™ refines that estimate over the recovery window in a way that no admission-only score can.

The neuroworsening results illustrate this distinction. Robertson’s earlier total-GFAP paper showed that the high trajectory group had delayed hematoma in 69.2% together with a right-shifted worst-Marshall CT distribution, indicating strong descriptive coupling between serial GFAP and hemorrhagic evolution [7]. Because that study used a latent trajectory-class framework and then related group membership to ICU-burden measures, it was optimized to describe serial phenotypes and their biological context rather than to test out-of-fold predictive discrimination. In our current reanalysis, the analogous descriptive signal was weaker but still directionally present: the high early neoGFAP™ tertile was enriched for more severe Marshall worsening and more delayed hematoma than the low tertile, yet cross-validated prediction of CT worsening and delayed hematoma remained poor. The exploratory latent-class analogue in the Supplementary Appendix reached a similar conclusion. It showed only partial structural resemblance to the Robertson SIMOA classes, with neoGFAP™ separating unfavorable outcome and ICU-burden measures somewhat more coherently than MSD total GFAP but without reproducing the published mortality gradient. These findings are not contradictory. Rather, they suggest that a biomarker can track hemorrhagic-complication burden descriptively without functioning as a robust stand-alone predictor.

### Limitations

Several limitations should remain explicit. Healthy-control data from a large reference cohort were not available for neoGFAP™; therefore, future work should consider reference-band or z-score approaches analogous to recent serum GFAP reference-interval and reference-database studies [9,10]. NfL was not measured in this cohort due to sample volume limitations, restricting our within-cohort platform comparison to total GFAP and neoGFAP™; prospective work incorporating NfL alongside neoGFAP™ is a natural extension, and the CENTER-TBI companion analysis on the Thermo ProQuantum™ platform provides an initial 3-BB (neoGFAP™, total GFAP, NfL) benchmark. Prospective validation should also test clinically interpretable trajectory features such as delta from baseline, early slope, time-to-peak, and area under the curve, because physician use is likely to depend more on within-patient change over time than on a single cut-point. The parent dataset did not include paired subject-level total-GFAP measurements on both SIMOA and MSD platforms, so a true individual-level MSD-versus-SIMOA total-GFAP correlation could not be calculated. The latent-class comparison can therefore be interpreted only as a structural or directional comparison, not as demonstrated cross-platform equivalence. Future studies should explicitly investigate cross-platform comparability and calibration harmonization between MSD and SIMOA, ideally using paired samples, shared calibrators, and prospective bridging cohorts. The single-site cohort was relatively small (n = 95), and the findings require validation in larger, independent, prospectively collected cohorts. Because this is a secondary analysis of archived data, findings should be interpreted as hypothesis-generating rather than definitive for clinical deployment.

Nonetheless, the results support further prospective evaluation of neoGFAP™ as a pathophysiologyinformed, proteoform-resolved astroglial monitoring biomarker after msTBI.

## Declarations

### Ethical Oversight

De-identified archived samples were obtained from previous observational studies approved by the Institutional Review Board at Baylor College of Medicine (protocol H-44131). That Institutional Review Board oversight covered the entire study, including all patient cohorts and all human samples and human data reported here.

### Informed Consent

Written informed consent or waiver of consent was obtained for all subjects per IRB-approved protocols.

### Consent for publication

Not applicable (de-identified data only).

### Data Availability

De-identified data from this study are available upon reasonable request to the corresponding author, subject to institutional data-sharing policies and patient privacy protections.

### Competing interests

WEH holds dual leadership roles and equity interest in Gryphon Bio and Owl Therapeutics. Other authors declare no competing interests.

### Funding

This work was supported by Gryphon Bio and by the Department of Defense under Awards W81XWH2110469 and HT94252310392 to Gryphon Bio. Opinions, interpretations, conclusions and recommendations are those of the authors and are not necessarily endorsed by the Department of Defense.

### Clinical Trial Registration

This study was a retrospective observational secondary analysis of archived samples and clinical data from previous observational studies; no intervention was prospectively assigned to participants. Clinical trial registration therefore does not apply.

### Authors’ contributions

KKW, ST, CR, and WEH conceptualized the work. KKW and WEH developed the methodology. FK, GC, KB, EE, DJ, and KT led the analysis of biomarkers in the deidentified samples. KKW and WEH performed the formal data analysis. KKW and WEH led manuscript writing with assistance from ST and CR. KR provided project administration and regulatory coordination. All authors read and approved the final manuscript.

## Acknowledgments

The authors thank the patients and families who participated in the original observational studies. We thank the clinical research coordinators and laboratory personnel who collected and processed samples. We thank Dr. Andrew Maas for critically reviewing this manuscript.

## Notes

### Author Declarations

De-identified archived samples were obtained from previous observational studies approved by the Institutional Review Board at Baylor College of Medicine (protocol H-44131). That Institutional Review Board oversight covered the entire study, including all patient cohorts and all human samples and human data reported here. This study was a retrospective observational secondary analysis of archived samples and clinical data from previous observational studies; no intervention was prospectively assigned to participants.

