## Supplemental for "Serial neoGFAP^™^ outperforms total GFAP for monitoring and 6-month outcome discrimination after moderate to severe traumatic brain injury: an exploratory single-site cohort study"

### S1. Extended Methods

This supplement contains extended methodology, secondary-endpoint model results, trajectory-based analyses, and the exploratory Robertson-style latent-class analogue. All analyses use the same MSD-platform serial data (n=95 patients, 651 draw-level measurements) described in Methods. NfL was not measured in this cohort due to sample volume limitations. Age-standardized $\log_{10}$ biomarker transformations are used where indicated. All AUROCs are reported to two decimal places.

### S2. Secondary endpoint cross-validated model performance

***Supplementary Table S2.*** *Full cross-validated performance across secondary endpoints (DRS ≥15, mortality, CT worsening, delayed hematoma, hemorrhagic composite).*

| Endpoint | Model | N | Events | AUROC | AUPRC | Brier |
|---|---|---|---|---|---|---|
| DRS ≥15 | Age + admission GCS | 89 | 25 | 0.73 | 0.51 | 0.19 |
| DRS ≥15 | IMPACT + early neoGFAP™ | 85 | 24 | 0.83 | 0.66 | 0.15 |
| DRS ≥15 | IMPACT + late neoGFAP™ | 82 | 23 | 0.84 | 0.68 | 0.14 |
| 6-month mortality | Age + admission GCS | 90 | 14 | 0.78 | 0.42 | 0.11 |
| 6-month mortality | IMPACT + early neoGFAP™ | 86 | 13 | 0.79 | 0.44 | 0.11 |
| 6-month mortality | IMPACT + late neoGFAP™ | 84 | 13 | 0.81 | 0.46 | 0.10 |
| CT worsening (≥+2) | Age + admission GCS | 95 | 15 | 0.57 | 0.24 | 0.14 |
| CT worsening (≥+2) | IMPACT + early neoGFAP™ | 91 | 14 | 0.61 | 0.28 | 0.13 |
| Delayed hematoma | Age + admission GCS | 95 | 30 | 0.55 | 0.35 | 0.22 |
| Delayed hematoma | IMPACT + early neoGFAP™ | 91 | 29 | 0.59 | 0.42 | 0.21 |
| Hemorrhagic composite | IMPACT benchmark | 95 | 37 | 0.65 | 0.51 | 0.21 |
| Hemorrhagic composite | IMPACT + early neoGFAP™ | 91 | 36 | 0.68 | 0.55 | 0.20 |

### S3. Trajectory-derived features (delta from baseline, slope, area-under-curve)

Trajectory summaries were computed per subject across the 6–240 h window. Baseline was the 6–24 h window mean; delta-from-baseline was computed at each subsequent sample. Whole-window slope was fit by ordinary least squares on $\log_{10}$ concentration versus hours. Area-under-the-curve was computed by trapezoidal integration on native-scale values. Neither delta, slope, nor AUC provided meaningfully improved cross-validated AUROC beyond the late-window mean captured in the main text Table 3.

### S4. Marshall CT worsening breakdown by early neoGFAP™ tertile

***Supplementary Table S7.*** *Descriptive Marshall CT worsening and delayed hematoma frequencies by early neoGFAP™ tertile.*

| Tertile | N | Δ Marshall ≥+2 | Delayed hematoma | Day-1 delayed hem. |
|---|---|---|---|---|
| Low | 30 | 1/30 (3.3%) | 7/30 (23.3%) | 5/7 (71%) |
| Mid | 30 | 3/30 (10.0%) | 11/30 (36.7%) | 6/11 (55%) |
| High | 30 | 5/30 (16.7%) | 12/30 (40.0%) | 8/12 (67%) |

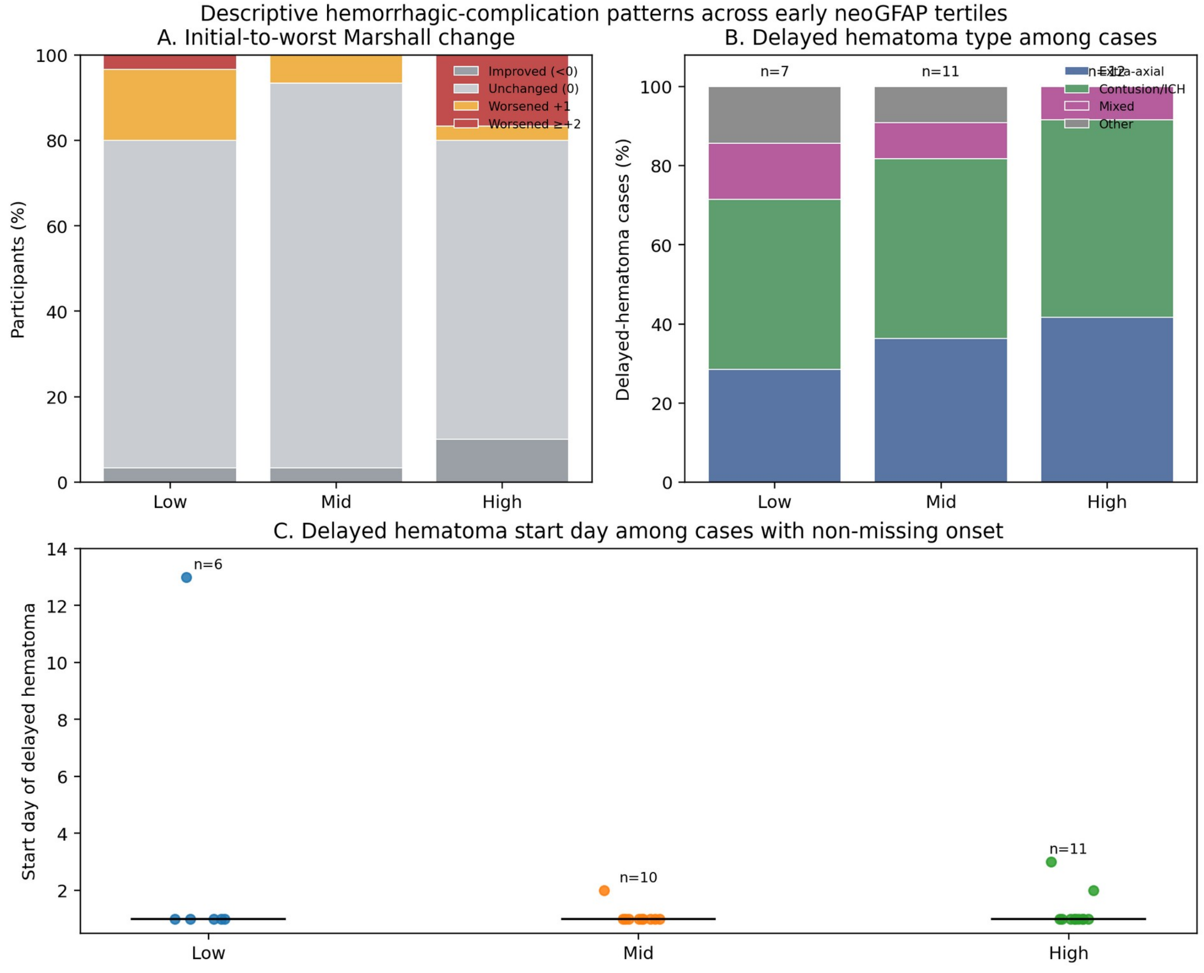


***Supplementary Figure S3.*** *Descriptive hemorrhagic-complication patterns across early neoGFAP™ tertiles. Panel A shows initial-to-worst Marshall CT code change grouped as improved (<0), unchanged (0), worsened +1, or worsened ≥+2. Panel B shows delayed hematoma type among delayed-hematoma cases grouped as extra-axial (SDH/EDH), contusion/ICH, mixed hemorrhagic pattern, or other/unspecified. Panel C shows delayed hematoma start day among cases with non-missing onset. Consistent with Supplementary Table S7, more severe Marshall worsening clustered in the high early neoGFAP™ tertile, whereas delayed hematoma onset was concentrated on day 1 across tertiles. In-figure labels use the earlier analyte name for this analyte.*

### S5. Exploratory Robertson-style latent-class analogue

To bridge our regression framework to Robertson's published TRAJ latent-class approach [7], we ran an exploratory Gaussian mixture on daily-binned $\log_{10}$ MSD serial data for both total GFAP and neoGFAP™. Both analytes preferred four classes by BIC, but three-class solutions retained posterior probability ≥0.85 for the majority of subjects and were used for apples-to-apples comparison with the Robertson 3-class model. MSD total GFAP partially resembled the SIMOA high-class result for unfavorable outcome (87.5% high class), while neoGFAP™ produced somewhat cleaner class separation for less-than-good recovery and ICU-burden measures. Neither analyte reproduced the strong mortality gradient reported for the SIMOA cohort, likely reflecting platform, sampling-window, and cohort-composition differences.

Supplementary Table S8 comprises panels S8A (model search) and S8B (three-class structural comparison). Model search considered K=2–4 classes and used BIC together with average posterior probability as selection criteria. Both MSD analytes preferred four classes by BIC, but the three-class solutions retained high average posterior probabilities and were therefore used for the clearest structural comparison with the published three-class Simoa GFAP model.

***Supplementary Table S8A.*** *Model-search summary for the exploratory Robertson-style latent-class analogue.*

| Biomarker | Classes (K) | BIC | Lowest avg posterior | Class sizes |
|---|---|---|---|---|
| MSD total GFAP | 2 | 906.2 | 0.941 | 41 / 50 |
| MSD total GFAP | 3 | 835.1 | 0.923 | 36 / 47 / 8 |
| MSD total GFAP | 4 | 796.7 | 0.917 | 8 / 42 / 26 / 15 |
| MSD neoGFAP™ | 2 | 919.2 | 0.930 | 58 / 33 |
| MSD neoGFAP™ | 3 | 820.2 | 0.902 | 18 / 42 / 31 |
| MSD neoGFAP™ | 4 | 812.9 | 0.833 | 18 / 24 / 18 / 31 |

Lower BIC indicates better fit. For both MSD total GFAP and MSD neoGFAP™, four classes gave the lowest BIC; the three-class solutions were retained for direct descriptive comparison with the published three-class Simoa GFAP model.

***Supplementary Table S8B.*** *Three-class structural comparison between the published Simoa GFAP model and the exploratory MSD latent-class analogues.*

| Study / analyte | Class sizes (L / M / H) | Poor GOSE % (L / M / H) | Mortality % (L / M / H) | Median average ICP mm Hg (L / M / H) |
|---|---|---|---|---|
| Robertson Simoa GFAP (published) | 23 / 48 / 26 | 30.4 / 54.2 / 88.5 | 8.7 / 6.3 / 26.9 | 14.4 / 15.9 / 17.6 |
| MSD total GFAP (3-class analogue) | 36 / 47 / 8 | 50.0 / 69.6 / 87.5 | 9.4 / 17.4 / 12.5 | 14.6 / 16.4 / 16.3 |
| MSD neoGFAP™ (3-class analogue) | 31 / 42 / 18 | 46.4 / 67.5 / 83.3 | 7.1 / 17.5 / 16.7 | 14.0 / 17.0 / 16.2 |

Poor GOSE denotes a 6-month GOSE of 4 or less. For poor GOSE, MSD total GFAP partly resembled the published Simoa total-GFAP pattern in the high class, whereas MSD neoGFAP™ showed somewhat cleaner separation across low, middle, and high classes. Neither MSD analyte reproduced the strong high-class mortality gradient reported in the published Simoa study.

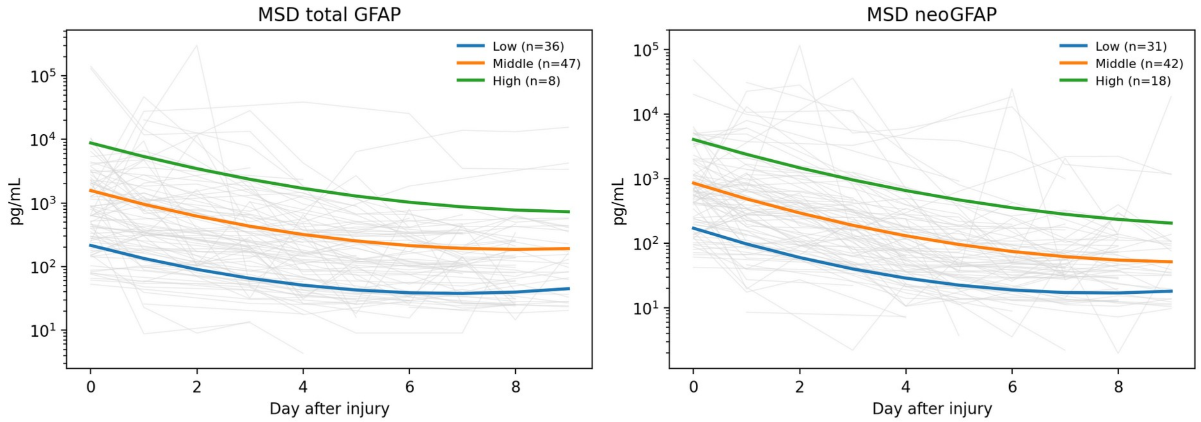


***Supplementary Figure S4.*** *Exploratory daily latent-class trajectory curves for MSD total GFAP and MSD neoGFAP™. Gray lines show individual daily-binned trajectories; colored lines show the three-class solutions retained for comparison with the published three-class Simoa model, even though BIC favored four classes for both analytes. neoGFAP™ yielded the more balanced three-class solution, whereas total GFAP concentrated the highest-risk class into a smaller subgroup. In-figure labels use the earlier analyte name for this analyte.*

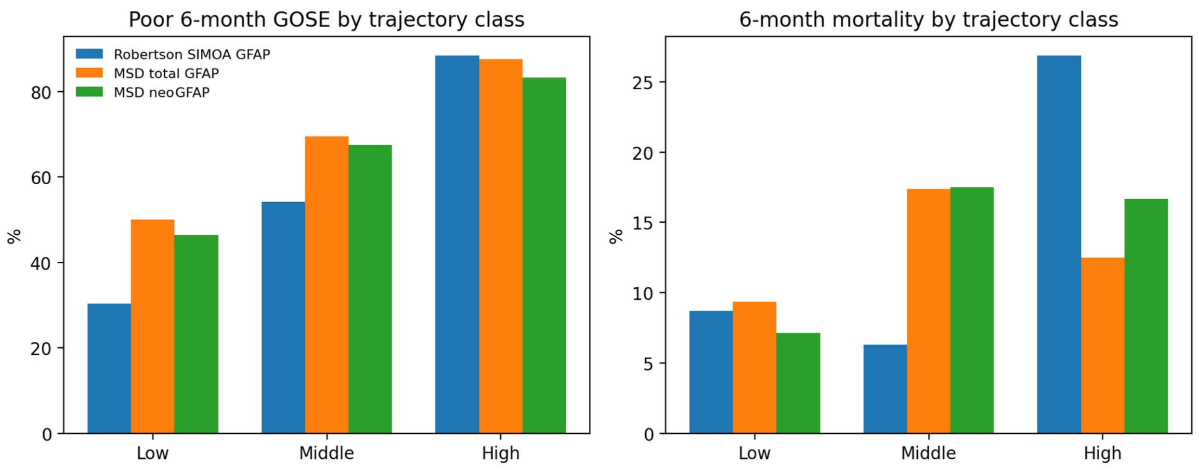


***Supplementary Figure S5.*** *Structural comparison of published Simoa GFAP trajectory classes with exploratory MSD total GFAP and MSD neoGFAP™ classes. Left, poor 6-month GOSE by trajectory class; right, 6-month mortality by trajectory class. Poor-GOSE gradients remain directionally similar across analytes, but the strong high-class mortality signal of the published Simoa study was not reproduced. In-figure labels use the earlier analyte name for this analyte.*
